# ANALYSIS OF SARS-COV-2 GENOME SEQUENCES FROM THE PHILIPPINES: GENETIC SURVEILLANCE AND TRANSMISSION DYNAMICS

**DOI:** 10.1101/2020.08.22.20180034

**Authors:** Francis A. Tablizo, Carlo M. Lapid, Benedict A. Maralit, Jan Michael C. Yap, Raul V. Destura, Marissa M. Alejandria, Joy Ann Petronio-Santos, El King D. Morado, Joshua Gregor A. Dizon, Jo-Hannah S. Llames, Shiela Mae M. Araiza, Kris P. Punayan, John Mark S. Velasco, Julius Aaron Mejia, Maribell Dollete, Sonia Salamat, Christina Tan, Kristianne Arielle D. Gabriel, Shebna Rose D. Fabilloren, Bernard Demot, Shana F. Genavia, Jarvin E. Nipales, Alessandra C. Sanchez, Haifa L. Gaza, Geraldine M. Arevalo, Coleen M. Pangilinan, Shaira A. Acosta, Melanie V. Salinas, Brian E. Schwem, Angelo D. Dela Tonga, Ma. Jowina H. Galarion, Niña Theresa P. Dungca, Stessi G. Geganzo, Neil Andrew D. Bascos, Eva Maria Cutiongco-de la Paz, Cynthia P. Saloma

**Affiliations:** Core Facility for Bioinformatics, Philippine Genome Center, University of the Philippines; DNA Sequencing Core Facility, Philippine Genome Center, University of the Philippines; Philippine Genome Center, University of the Philippines; National Institutes of Health, University of the Philippines Manila; Institute of Clinical Epidemiology, National Institutes of Health, University of the Philippines Manila; Division of Infectious Diseases, Department of Medicine, University of the Philippines Philippine General Hospital; Natural Sciences Research Institute, College of Science, University of the Philippines Diliman; National Institute of Molecular Biology and Biotechnology, University of the Philippines Diliman

**Keywords:** SARS-CoV-2, COVID-19, Philippines, genetic surveillance, community transmission

## Abstract

The spread of the corona virus around the world has spurred travel restrictions and community lockdowns to manage the transmission of infection. In the Philippines, with a large population of overseas Filipino contract workers (OFWs), as well as foreign workers in the local online gaming industry and visitors from nearby countries, the first reported cases were from a Chinese couple visiting the country in mid-January 2020. Three months on, by mid-March, the COVID-19 cases in the Philippines had reached its first 100, before it exploded to the present 178,022 cases (as of August 20, 2020). Here, we report a genomic survey of six (6) whole genomes of the SARS-CoV-2 virus collected from COVID-19 patients seen at the Philippine General Hospital, the major referral hospital for COVID-19 cases in Metro Manila at about the time the Philippines had over a hundred cases. Analysis of commonly observed variants did not reveal a clear pattern of the virus evolving towards a more infectious and severe strain. When combined with other available viral sequences from the Philippines and from GISAID, phylogenomic analysis reveal that the sequenced Philippine isolates can be classified into three primary groups based on collection dates and possible infection sources: (1) January samples collected in the early phases of the pandemic that are closely associated with isolates from Wuhan, China; (2) March samples that are mainly linked to the M/V Diamond Princess Cruise Ship outbreak; and (3) June samples that clustered with European isolates, one of which already harbor the globally prevalent D614G mutation which initially circulated in Europe. The presence of community-acquired viral transmission amidst compulsory and strict quarantine protocols, particularly for repatriated Filipino workers, highlights the need for a refinement of the quarantine, testing, and tracing strategies currently being implemented to adapt to the current pandemic situation.

## INTRODUCTION

The COVID-19 pandemic has spread to 213 countries and territories from the original epicenter of the outbreak in Wuhan, China. The first two confirmed cases of COVID-19 in the Philippines were tourists, a Chinese couple from Wuhan, China. They developed symptoms for COVID-19 on January 18 and 21, and got the first laboratory test confirmation on January 30. The earliest reported case of local transmission involved a Filipino couple who were hospitalized in the first week of March (Coronavirus disease (COVID-19) Situation Report 2 Philippines 11 March 2020, WHO). By March 11, the Philippines had 49 confirmed cases with 31% of those being imported cases from China, Japan, South Korea, Australia, UAE as well as from the M/V Diamond Princess Cruise Ship being quarantined in Yokohama, Japan (World Health Organization 2020, March 9 and 11). The reported number of SARS-CoV-2 positive cases in the Philippines reached its first 100 in mid-March.

The Philippines attracts a large number of regional tourists coming from China and South Korea, which have earlier been reported to have large outbreaks of COVID-19. As a consequence, the Philippines, similar to many countries around the region, limited and finally closed air travel first from these regions. The return of overseas Filipino workers (OFWs) comprising a large population of those engaged in the shipping and cruise ship industries which have been hard-hit by COVID-19 infections, necessitated a policy of testing returning OFWs through rapid antibody-based testing and by RT-PCR, as well as a 14-day quarantine or self-isolation before they are allowed to travel to their onward destinations in the various cities and provinces of the Philippines.

To control the spread of the virus, the government placed the entire island of Luzon under enhanced community quarantine (ECQ) from 17 March to 31 May, 2020 prohibiting air and sea travel into and out of the whole island of 63 million people with restrictions in land travel within. By this time, the spread of the virus in the country has been through community infection.

In this study, we utilized shotgun metagenomic sequencing of samples collected on 22 to 26 March 2020 from COVID-19 positive cases who have been detected in the course of the field validation of a locally-developed RT-PCR based SARS-CoV-2 detection kit, the GenAmplify nCoV rRT-PCR test kit. These patients were admitted at the Philippine General Hospital, a major COVID-19 referral hospital in the metropolis. We report the detection of a total of 48 variants, five of which are common, across all six Philippine isolates sequenced in this study compared to the Wuhan reference sequence. By combining our data with other available SARS-CoV-2 sequences from the Philippines and the GISAID database, we present some insights on the transmission dynamics of the strains of SARS-CoV-2 circulating in the country, as well as their possible implications on current COVID-19 containment strategies.

## METHODOLOGY

### Ethics Approval

Study participants were enrolled as part of the field validation trial of a locally developed RT-PCR detection kit, the GenAmplify COVID-19 rRT-PCR Detection Kit. The study received ethics approval from the University of the Philippines Manila Research Ethics Board with approval number 2020-187-01.

### Sample Collection and Viral RNA Extraction

Nasopharyngeal and/or oropharyngeal swabs were collected from patients between March 22 to March 28, 2020. The collected samples were then subjected to heat inactivation and transported in viral transport media. Upon arrival, the inactivated samples were centrifuged for 10 min. at 1500 × g to separate non-viral cells and minimize DNA co-purification. A 140 uL volume of the resulting supernatant was then processed for RNA extraction using QIAamp Viral RNA Mini Kit (Qiagen), following the manufacturer’s protocol except for the addition of carrier RNA to minimize the occurrence of polyA-tailed sequencing reads. The quantity and purity of the extracted RNA samples were measured using Qubit HS RNA assay (Invitrogen) and NanoDrop8000 (Thermo Fisher Scientific), respectively.

### Library Preparation and Sequencing

An input amount of 50 ng to 100 ng of RNA extract was used to generate 260-300bp sequencing libraries using the TruSeq Total RNA H/M/R Library Preparation Kit (Illumina). Quantification of the libraries was done using Qubit HS dsDNA assay (Invitrogen) and the library sizes were determined using TapeStation 2200 (Agilent). All of the libraries were normalized to a concentration of 4nM prior to pooling. Final dilutions of 1.5 pM pooled libraries were then loaded for sequencing in NextSeq 550 (2×150 bp PE) using the NextSeq 500/550 Mid-Output Kit v2.5 (Illumina). Samples were multiplexed to have at least 10 million sequencing reads per sample and were subsequently demultiplexed using bcl2fastq v2.20.

### Sequence Quality Control and Filtering

Raw demultiplexed sequence data from each of the Philippine SARS-CoV-2 isolates were subjected to quality filtering using the tool fastp (Chen et al. 2018) with default parameters. All reads passing the initial quality control step were further filtered using two different and separate procedures: (1) the “*human-filtered*” procedure wherein the reads were initially mapped to the human hg38 reference genome and all unmapped reads were selected for subsequent meta-assembly and (2) the “*betacov-filtered*” procedure wherein reads were initially mapped to a database of Betacoronavirus sequences and those that mapped were used for subsequent meta-assembly. In both of the described procedures, the tool BWA (Li and Durbin 2009) was used for mapping followed by filtering and conversion from BAM to FASTQ format using Samtools (Li et al. 2009).

### Assembly and Annotation of the SARS-CoV-2 Genomes

All remaining reads after the filtering steps were assembled using metaSPAdes (Nurk et al. 2017). Note that separate assemblies were made for the human- and betacov-filtered reads. The resulting contigs were then matched against a database of SARS-CoV-2 genome sequences using BLAST (Altschul et al. 1990), and those with significant matches (E-value of 1×10^−3^ or less; query coverage of at least 50%) were collected. From this subset, we then compared the human- and betacov-filtered assemblies for each sample, selecting those with a total size of > 29 Kb, longer contig lengths, and fewer overall contigs as the better assembly.

The chosen assemblies were further refined by scaffolding based on BLAST alignment coordinates against the NCBI reference SARS-CoV-2 sequence (NC_045512.2) using a custom Python script. Briefly, contigs were arranged based on their mapping coordinates. Contigs with overlapping coordinates were collapsed, and regions without coverage were filled with “N”. The resulting scaffolds were then annotated using RATT (Otto et al. 2011) and VAPiD (Shean et al. 2019). The overall sequence and structural similarities of the scaffolds with the aforementioned reference sequence were also observed using MAUVE (Darling et al. 2004). Nearly complete scaffolds were then deposited in the EpiCoV database of the Global Initiative on Sharing All Influenza Data (GISAID) (Elbe and Buckland-Merrett 2017; Shu and McCauley 2017).

### Variant Analysis and Gene Alignments

Variants were obtained from the output of MUMmer (Kurtz et al. 2004), implemented as part of the RATT annotation transfer workflow. The MUMmer SNP output was converted to VCF format using a simple script written in Python, and the VCF files were used as input to snpEff (Cingolani et al. 2012) for variant annotation.

For surveillance purposes, nucleotide and amino acid sequences for five critical SARS-CoV-2 protein products (RNA-dependent RNA polymerase, spike glycoprotein, membrane glycoprotein, envelope protein, and the nucleocapsid phosphoprotein), were extracted from the annotated scaffolds and aligned using MAFFT (Katoh et al. 2002). Manual adjustments to the alignments were made as needed using BioEdit (Hall 1999) to correct for errors in annotation transfer. Alignments were then viewed using MView (Brown, Leroy, and Sander 1998).

Possible structural and functional consequences of the more commonly observed variants were also inferred based on protein structure models generated via the C-I-TASSER pipeline (Zheng et al. 2019), obtained from I-TASSER’s COVID-19 website (https://zhanglab.ccmb.med.umich.edu/COVID-19/).

### Phylogenomic Analysis

An initial maximum likelihood tree from 246 complete SARS-COV-2 genome sequences obtained from the GISAID EpiCoV database (https://www.gisaid.org/; accessed on March 09, 2020) was generated by first aligning the sequences using MAFFT. The resulting alignment was then trimmed using TrimAl (Capella-Gutiérrez, Silla-Martínez, and Gabaldón 2009). The tool jModelTest2 (Darriba et al. 2012) was used to determine the best nucleotide substitution model for the trimmed alignment. Maximum likelihood tree reconstruction was finally implemented using RAxML (Stamatakis 2014) with the GTRGAMMA model (as determined via jModelTest2) and 1000 bootstraps for node support.

The six scaffold assemblies from the Philippine isolates, together with 1,083 SARS-COV-2 genome sequences also from GISAID (accessed on March 26, 2020, May 26, 2020, and August 06, 2020), were added to the initial alignment of 246 sequences using MAFFT and subsequently trimmed with TrimAl. The evolutionary placement algorithm implemented using RAxML was then used to determine the phylogenetic positions of all the additional isolates using the previously generated maximum likelihood tree as seed tree. The resulting tree was then visualized and annotated primarily using the R ggtree package (Yu et al. 2017).

## RESULTS

### Community acquired infections among COVID-19 patients in Metro Manila in March 2020

At the time the samples were collected between March 22 to 26, 2020, the entire island of Luzon, where the National Capital Region (est. population in 2020 at 13.9 million) is situated, was already placed under enhanced community quarantine (ECQ) –- a situation where land, sea and air travel were not permitted. All six (6) patients are from Metro Manila and all have had no travel history outside the country on the month before contracting the SARS-CoV-2 virus. Two patients had close contact with a confirmed case of COVID-19, one of whom gave direct care to a hospitalized relative while the other was a physician whose wife was exposed to a confirmed case. Both patients had no comorbid conditions and presented with mild symptoms of fever plus dry cough, myalgia, headache and diarrhea. Four patients had severe COVID-19 pneumonia, two with exposure to suspect cases and two with no known exposure to a confirmed or probable case of COVID-19. Two patients needed mechanical ventilation (**Table 1**).

**Table 1.** Profile of COVID-19 Patients. All six (6) patients came from Metro Manila and all have had no travel history outside the country on the month before contracting the SARS-CoV-2 virus. Their symptoms range from mild fever to severe with dyspnea and epigastric pain in one case.

| PGC Code | Date of Collection | History/ contact tracing | Severity of illness | Outcome | Symptoms |
| --- | --- | --- | --- | --- | --- |
| PGC001 | 3/22/2020 | 42yo male; no travel history, works as a private car hire driver with close contact to COVID-19 probable individual | Severe | Recovered | Fever, cough, sore throat, chills |
| PGC002 | 3-26-2020 | 33yo male exposed to a COVID-19 confirmed case | Mild | Recovered | Fever |
| PGC003 | 3-26-2020 | 56yo male with no travel history and no known exposure to a COVID-19 case. | Severe | Died 4/7/20 | Cough, fever, sore throat, dyspnea |
| PGC004 | 3/26/2020 | 28yo male health care worker (physician) exposed to a confirmed case | Mild | Recovered | Fever, headache, cough, myalgia |
| PGC005 | 3/27/2020 | 82yo female from Manila, no travel history, bed-bound, with exposure to COVID-19 probable individuals (symptomatic household members) | Severe | Died 3/28/20 | Fever, dyspnea, somnolence |
| PGC006 | 3/28/2020 | 42yo male with no known exposure to a COVID-19 case | Severe | Recovered | Cough, dyspnea, epigastric pain, nausea, vomiting |

Two of the four patients with severe pneumonia died due to progression of pneumonia to acute respiratory distress syndrome (ARDS). Both patients were bed-bound, one of whom was an elderly woman with no other illnesses but with history of exposure to household members with symptoms of probable COVID-19; while the other one had a pre-existing ischemic stroke with no known exposure to a confirmed or probable case. Both of the patients with severe pneumonia who survived were in their early 40s, one with no comorbid condition while the other had stable hypertension. Of the six patients, the four who survived recovered with no symptoms, despite prolonged viral shedding ranging from six to seven weeks.

### SARS-CoV-2 Genome Assembly and Annotation

In this study, nearly complete genome scaffolds for six Philippine SARS-CoV-2 samples were generated and are now publicly available through the EpiCoV database of GISAID (**Table 2**). These scaffolds were found to be highly similar with the reference SARS-CoV-2 genome from NCBI GenBank (NC_045512.2) in terms of sequence and organization (**Figures S1**). All genomes were predicted to harbor 11 genes classically arranged in the following order: ORF1ab polyprotein – Spike glycoprotein (S) – ORF3a – Envelope protein (E) – Membrane glycoprotein (M) – ORF6 – ORF7a – ORF7b – ORF8 – Nucleocapsid phosphoprotein (N) – ORF10.

**Table 2.** Assembly statistics from the genome scaffolds of the six Philippine SARS-CoV-2 isolates. The GISAID accession IDs and the number of predicted variant positions relative to the NCBI GenBank NC_045512.2 reference sequence are also shown.

| Sample Code | Scaffold Length (bp) | Potential Coverage | % Ambiguous Bases (% N's) | GISAID Accession ID | # Predicted Variants (Unique to Sample) |
| --- | --- | --- | --- | --- | --- |
| PGC001 | 29,871 | 260x | 2.37% | EPI_ISL_431833 | 14 (8) |
| PGC002 | 29,981 | 177x | 0.19% | EPI_ISL_434554 | 14 (7) |
| PGC003 | 29,869 | 498x | 0.04% | EPI_ISL_434555 | 9 (1) |
| PGC004 | 29,873 | 99x | 1.16% | EPI_ISL_434556 | 14 (7) |
| PGC005 | 29,871 | 27x | 2.53% | EPI_ISL_434557 | 16 (10) |
| PGC006 | 29,869 | 455x | 0.04% | EPI_ISL_434558 | 6 (0) |

### Gene Alignments

To facilitate genetic surveillance efforts, we looked at sequence alignments for five critical protein products of the SARS-CoV-2 genome (**Figures S2-S6**). Mutations in the RdRp, S, M, E, and N protein products may have considerable effects on current diagnostic and vaccine design efforts (Yong, Su, and Yang 2020). Among these genes, the envelope protein was found to be the most conserved, with 100% sequence similarity at both nucleotide and amino acid levels relative to the reference. The lowest sequence similarity was observed in the M gene of PGC001, with protein 91.1% sequence identity, due to a stretch of ambiguous bases (‘N’s’) in the scaffold assembly for that sample. A 35-bp repeat was also observed for this gene in the underlying nucleotide assembly of sample PGC002 (**data not shown**), resulting in a stretch of mismatched residues. All the other gene alignments revealed greater than 99% nucleotide and amino acid identities between the sequences of the Philippine samples and that of the reference.

### Variant Analysis

A total of 48 variant positions relative to the NC_045512.2 reference sequence were predicted across the six SARS-CoV-2 genome scaffolds. Among these variants, 14 have already been observed in at least one other SARS-CoV-2 genome deposited in the GISAID database, while five of these were found to be shared by all six isolates. Among the samples, the highest variant count (16 variants) was predicted for PGC005 (10 of which are unique to the sample), whereas the most similar to the reference was PGC006 with only six predicted variants, none of which were unique to the sample (**Table 3**).

**Table 3.** Variants common to all six Philippine SARS-CoV-2 samples. The variant positions shown are relative to the nucleotide positions in the NC_045512.2 reference sequence.

| Variant Position | Gene | Ref Allele | Alt Allele | Variant Type | Effect |
| --- | --- | --- | --- | --- | --- |
| 6,312 | ORF1ab (nsp3) | C | A | missense | T2016K (T1198K) |
| 11,083 | ORF1ab (nsp6) | G | T | missense | L3606F (L37F) |
| 13,730 | ORF1ab (RdRp) | C | T | missense | A4489V (A97V) |
| 23,929 | S | C | T | silent | Y789Y |
| 28,311 | N | C | T | missense | P13L |

The five variants shared by all six Philippine samples are listed in **Table 3** and their corresponding structural contexts are shown in **Figure 1**. Interestingly, these variants were found to occur more frequently in other SARS-CoV-2 genome sequences deposited in GISAID. In fact, all five of these variants were also observed in at least eleven other Philippine SARS-CoV-2 genome sequences in the GISAID database submitted by the Research Institute for Tropical Medicine (RITM), Department of Health, Philippines for clinical samples collected in March (**data not shown**). The only exceptions are for the L3606F amino acid replacement at in the ORF1ab gene, which was not found in RITM sample EPI_ISL_430456; and for RITM sample EPI_ISL_491470, for which ambiguous bases in the assembly prevent verification of three of the five variants.

**Figure 1.**
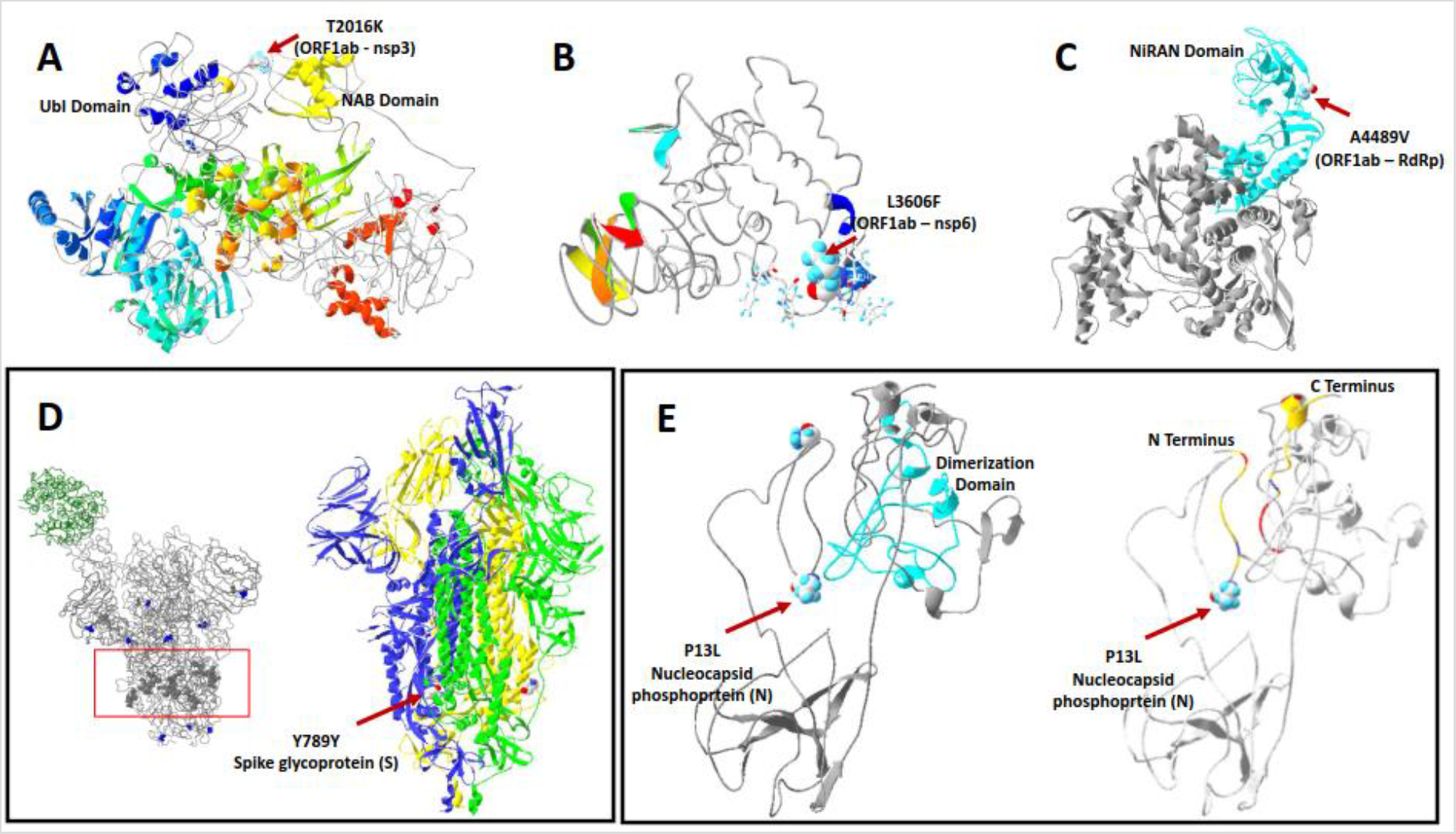
Structural analysis of the five variants shared by all six Philippine samples. The protein structure models were generated using the C-I-TASSER pipeline and directly obtained from the ITASSER COVID-19 website https://zhanglab.ccmb.med.umich.edu/COVID-19/. **(A)** Predicted structure of the NSP3 protein highlighting the T2016K mutation (red arrow). The UbI and NAB domains are also shown in blue and yellow, respectively. **(B)** Predicted structure of the NSP6 protein highlighting the L2606F mutation (red arrow). **(C)** Predicted structure of the RNA-dependent RNA-polymerase (RdRP) highlighting the A4498V mutation (red arrow). The NiRAN domain of the protein is also shown in cyan. **(D)** Left – Spike glycoprotein structure obtained from GISAID showing mutation hotspots (gray balls inside red box); Right – Predicted structure of the spike glycoprotein highlighting the silent mutation at amino acid position 789 (red arrow). **(E)** Predicted structure of the nucleocapsid phosphoprotein highlighting the P13L mutation (red arrow). Left – The dimerization domain of the nucleocapsid is shown in cyan; Right – The N- and C-terminus of the nucleocapsid are colored based on charge, with primarily acidic residues in red and basic in blue.

### Phylogenetic Analysis

The molecular phylogeny of 1,335 SARS-CoV-2 sequences was reconstructed based on whole genome sequence alignments. The observed clustering of the six samples from this study, as well as 17 other Philippine samples deposited in the GISAID database by RITM, revealed that these local isolates primarily grouped into eight clades (**Figure 2**). Nonetheless, in terms of hypothesized transmission, the isolates appear to have three primary sources: (1) early samples collected in January are closely related with isolates from Wuhan, China (**Figure 2, D and E**); (2) samples collected in March are primarily linked to the Diamond Princess Cruise ship outbreak (**Figure 2, G, H, and I**), with only one isolate clustering with viruses mainly from Shanghai, China (**Figure 2-F**); and (3) samples collected in June which are primarily associated with European isolates (**Figure 2, B and C**) We also note the detection of the D614G variant in one out of the two Philippine samples reportedly collected in June (EPI_ISL_491298).

**Figure 2.**
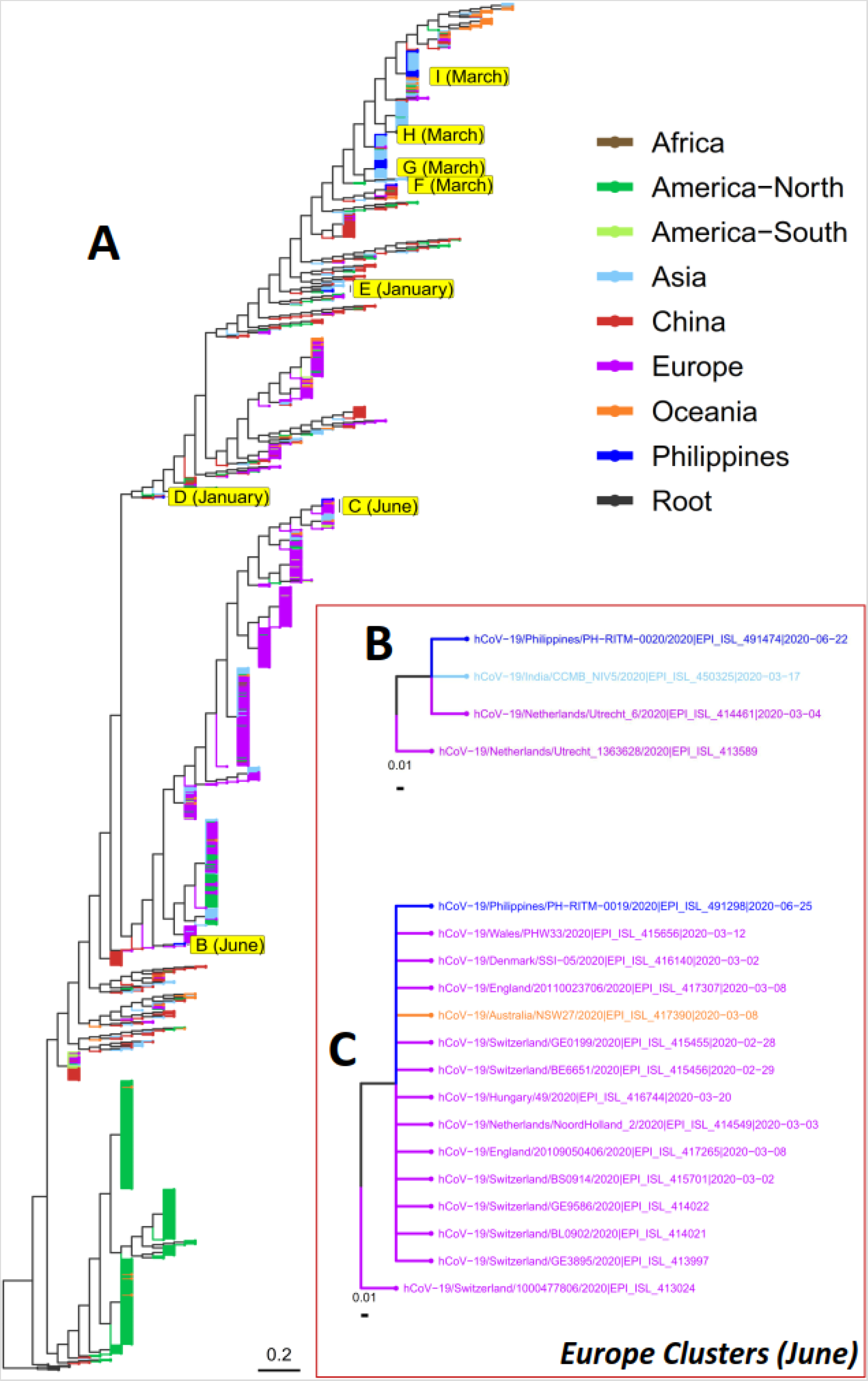
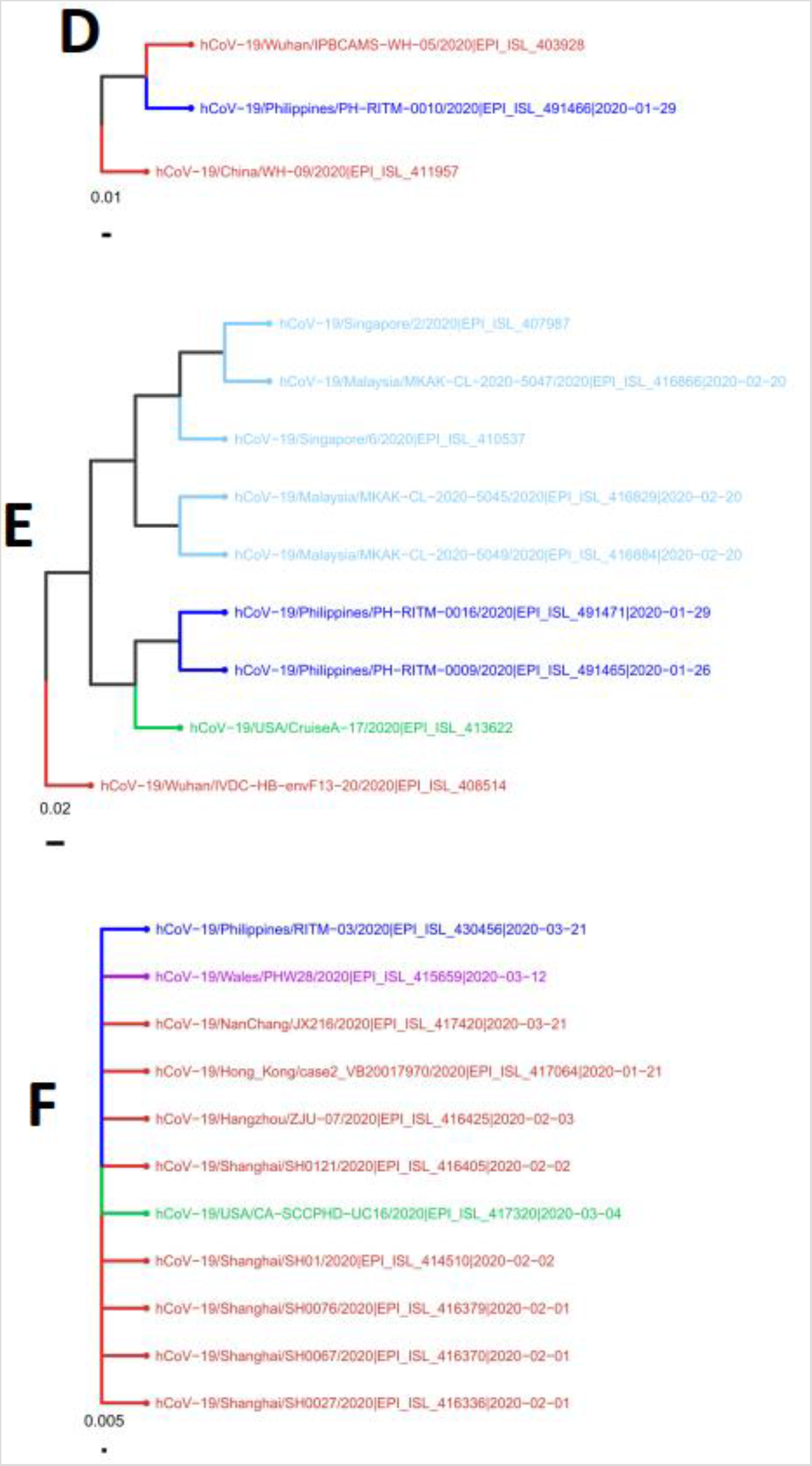
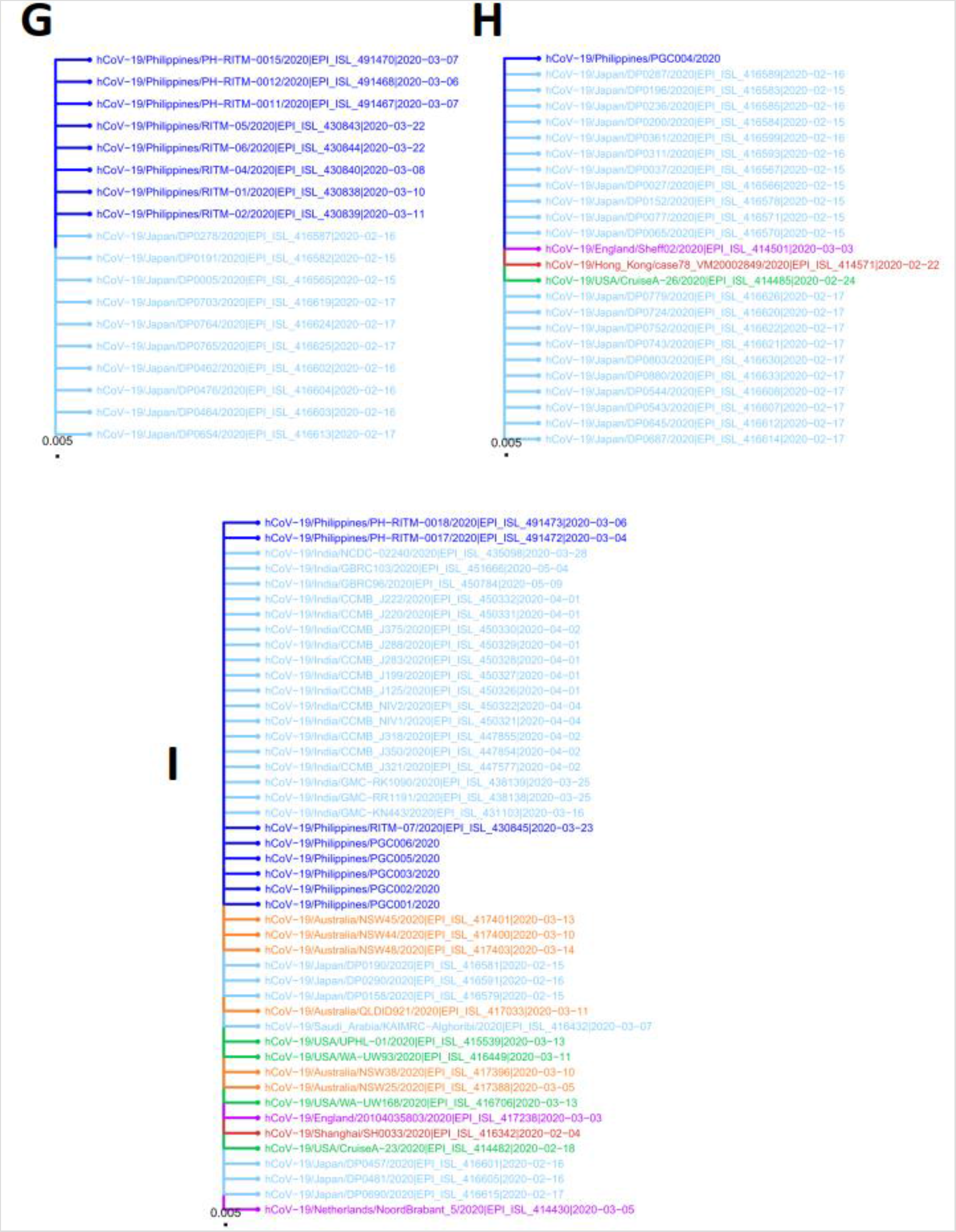
Phylogenetic analysis of SARS-CoV-2 genome sequences. The molecular phylogeny of 1,335 SARS-CoV-2 genome sequences is shown in **A**. The tree was reconstructed by phylogenetically placing a total of 1,089 sequences, including 1,083 from the GISAID database (17 of which were collected in the Philippines) and six local isolates from this study, to an initial maximum likelihood tree comprised of 246 sequences obtained earlier from GISAID (rooted with sequences from five pangolins and one bat). The tips were colored according to geographic locations, mostly corresponding to the continental origins of the isolates. For Asia however, two countries were categorized separately: China which is believed to be the origin of SARS-CoV-2 and the Philippines where the isolates from this study were collected. **B** and **C** are subtrees showing the clustering of Philippine isolates collected in June with samples mainly from Europe. **D, E**, and **F** are subtrees showing the clustering of Philippine isolates collected in January and March with samples mainly from China. **G, H**, and **I** are subtrees showing the clustering of Philippine isolates collected in March with samples linked to the M/V Diamond Princess Cruise Ship outbreak.

## DISCUSSION

We report here the full sequences of six (6) local SARS-CoV-2 isolates, all of which were collected within the month of March from patients in Metro Manila, Philippines. These patients had no travel history in foreign countries or in regions with active COVID-19 outbreaks, although some of them were engaged in activities that can increase the risk of infection (i.e., health care worker and private car-for-hire driver). By performing whole genome shotgun sequencing, it was possible to get insights into the circulating strains of the virus in the Philippines at this time and study critical regions in the viral genome for variations and mutations that will impact the RT-PCR kits being developed and utilized locally, as well as in understanding the spread and evolution of the virus.

### Genetic Surveillance

The nearly complete genomic scaffolds of six SARS-CoV-2 samples collected from the Philippines revealed that most of the variants observed were unique to a single isolate, indicating the possibility of rare, recent mutations or sequencing error. However, several variants were found to occur more frequently and are more likely to be true variants segregating at high frequency in the circulating viral population. Thus, these variants bear greater importance in evolutionary and genetic surveillance studies on the virus. For the Philippine samples, five variants were commonly observed: T2016K, L3606F and A4489V all within the ORF1ab gene; a silent mutation Y789Y in the S gene; and a P13L amino acid replacement found in the N gene (**Table 3**).

The T2016K variant at ORF1ab affects the region encoding for the NSP3 protein. In particular, this variant can be found between the UbI and NAB domains of the protein, both domains have been associated with nucleic acid binding (**Figure 1-A**). The Threonine to Lysine mutation increases the positive charge in the area, potentially increasing the nucleic acid binding affinity of NSP3.

The presence of the L3606F mutation (**Figure 1-B**) in SARS-CoV-2 genome has already been described previously (Benvenuto et al. 2020). This particular variation was found in the ORF1ab region encoding for the NSP6 protein. According to Benvenuto et al. (2020), this mutation increases the number of phenylalanines in the transmembrane domain of the said protein. This is hypothesized to generate a less flexible helix due to the stacking of aromatic rings, which may decrease overall protein stability. However, the increased aromaticity is inferred to also facilitate more stable binding with the endoplasmic reticulum and may eventually affect autophagosome formation.

The A4489V variant provides a conservative substitution within the NiRAN domain of the RdRp protein, which is also encoded by ORF1ab (**Figure 1-C**). While the altered residue is not located in the contacting surfaces of RdRp and NiRAN, its location between the subdomains in the NiRAN suggest a potential alteration of movement between these structures. The bulkier Valine residue is likely to provide less flexibility, potentially altering subdomain movement. This may affect the nucleotidyl transferase efficiency of the domain.

A silent mutation (Y789Y) was observed within the Spike glycoprotein (**Figure 1-D, Right**). This mutation occurs in a region observed to be highly variable in other genomic sequences (**Figure 1-D, Left**). Interestingly, the observed variation resulted in a codon that is similar to the one present in bat SARS-CoV-2. Based on human codon usage, the reference codon TAC (57%) is more preferred than the variant TAT (43%) observed in Philippine and bat samples. This change may represent a potential shift towards a less virulent strain, less deadly for the host, with greater probability of persistence.

The P13L variant exists at the turn from the initial strand (AA 1-13) to the next antiparallel strand (AA 14 – 23) of the nucleocapsid. These residues appear to interact with a surface of the dimerization domain, possibly aiding in attaining a more packed conformation (**Figure 1-E, Left**). Coloring the residues on the most N-terminal and most C-terminal strands (**Figure 1-E, Right**) reveal the presence of complementary charged residues that may aid their packing. The interaction of the N and C terminal strands also provide a “closed system” that stabilizes the protein structure. Alteration of P13 removes the kink, likely shifting the strand position that can destabilize the protein structure. Destabilization of the nucleocapsid may hinder viral particle production.

### Viral Transmission Dynamics

Phylogenomic analysis of 1,335 SARS-CoV-2 genome sequences (**Figure 2-A**), including 23 Philippine samples (six from this study), shows that samples from China are present at the base of every major clade – supporting the China origin of the virus. Later, localized community transmission can be observed, particularly for certain isolates from North America (Green) and Europe (Purple). Samples from Asia (Light Blue) and Oceania (Orange), including the Philippines (Blue), can be found throughout the tree, suggesting multiple points of viral entry in these regions.

The Philippine isolates were found to cluster into eight clades (**Figure 2, B to I**). Interestingly, these isolates can be further classified into three groups based on possible entry routes of the infection: (1) the China clusters of January samples, (2) the M/V Diamond Princess clusters of March samples, and (3) the Europe clusters of June samples.

Two samples collected in the month of June clustered mainly with European isolates (**Figure 2, B and C**), one of which carries the now globally prevalent D614G mutation that was initially observed to circulate more frequently in Europe. We note that the said variant was not detected in any of the locally sequenced SARS-CoV-2 isolates before June. However, a number of partially sequenced local samples collected in June and July were already found to harbor the mutation (**data not shown**). While the limited sampling warrants cautious interpretation, these observations suggest that the D614G mutation, albeit detected much later in the country, might also be increasing in occurrence following the globally observed trend.

Viral samples collected in January, during the early phases of the infection in the country, clustered with isolates from Wuhan, China – the epicenter of the pandemic at that time (**Figure 2, D and E**). This particular clustering was expected because the January samples were collected from Chinese nationals who traveled to the Philippines from Wuhan. We note that in Figure 2-E, the two Philippine isolates appear to group more closely with an isolate from the United States (EPI_ISL_413622). However, the USA isolate was reportedly collected on February 24, 2020, much later than the collection dates of the Philippine samples (January 26 and 29, 2020). In this context, we believe that the Philippine and USA samples were all linked to the Wuhan isolate (EPI_ISL_408514) which was reportedly collected on January 01, 2020.

Majority (18 out of 23) of the local samples with genome sequences were collected within the month of March. Among these samples, one clustered with isolates mainly coming from Shanghai, China (**Figure 2-F**). All the remaining samples were observed to group into clades mostly linked to the M/V Diamond Princess Cruise Ship outbreak (**Figure 2, H and I)**. Towards late February, passengers and crew members from various nationalities, including Filipinos, Indians, and Australians, were repatriated from the cruise ship. Notably, many of the Philippine isolates in these clusters (**Figure 2, H and I)** were sourced from individuals who had no travel history outside the country, suggesting the presence of community-acquired transmission possibly arising from undetected cases of infection in repatriated seafarers. Nonetheless, the strict imposition of the Enhanced Community Quarantine (ECQ) in the entire island of Luzon, where majority of the confirmed COVID-19 cases originated, from March 17 to May 31, 2020 might have stymied the further spread of the virus from the Diamond Princess cluster of cases, as no such related cases were observed in the months of June and July (cases of which are mostly linked to European clusters). We note, though, that these observations come from only a few local viral isolates with available sequence data and has a limited geographic reach as these were mostly collected in Metro Manila.

### Implications and Future Directions

Based on observations from the common variants, there is no clear pattern as to whether the SARS-CoV-2 genome is evolving towards a higher or lower virulence state. Nonetheless, most of the variants observed fall outside the target regions for viral diagnostics in the Philippines, suggesting that current testing procedures remain effective. Furthermore, the high sequence similarity among critical gene regions (RdRp, S, M, E, and N) suggest that diagnostic and vaccine design efforts are not considerably undermined by the presence of these mutations. However, these inferences are drawn from very limited samples, and more genomic and genetic data are necessary in order to provide a better understanding of the present evolutionary and genetic landscape of SARS-CoV-2 in the country.

Interestingly, all the source individuals of the six Philippine samples reported in this study had no travel histories outside the country and some without close contact with a known SARS-CoV-2 infected patient (**Table 1**). This body of information suggests the occurrence of community acquired infections and that a number of infected individuals remained undetected during the transmission period. Furthermore, the presence of undetected transmission reflect the challenges faced in implementing quarantine, testing, and tracing protocols in the country. A review of the current procedures may be warranted as the SARS-CoV-2 infection in the country continues to increase – with a substantial number of infections coming from repatriated overseas Filipino workers, as well as from locally stranded individuals traveling back to their home provinces.

## CONCLUSIONS

In this study, six nearly complete genome scaffolds of SARS-CoV-2 samples from the Philippines are reported. Variant analysis revealed the presence of five common variants that are most likely to be segregating at high frequency in the circulating local viral populations: T2016K, L3606F and A4489V all within the ORF1ab gene; a silent mutation Y789Y in the S gene; and a P13L amino acid replacement found in the N gene. Structural insights on these variants do not suggest that the the virus is shifting towards a more virulent and lethal strain. Transmission dynamics inferred from the phylogenomic clustering of the Philippine SARS-CoV-2 samples revealed three possible primary sources of the virus in the country: (1) early samples collected in January are closely related to isolates from Wuhan, China; (2) samples collected in March are mainly associated with the M/V Diamond Princess Cruise Ship outbreak; and (3) samples collected in June can be linked to isolates from Europe. Considering that many of the local isolates were collected from individuals without travel histories outside the country and some have no known interaction with a confirmed positive case, these findings highlight the need to further improve the current quarantine, testing, and tracing protocols being employed locally to adapt to the current pandemic situation. Even though no association can be made between the observed phylogenomic clustering and medical presentation, advanced age still appears to be a risk factor for disease severity – although co-morbidities can also substantially affect clinical outcomes.

## Data Availability

Genome sequences of the six Philippine SARS-CoV-2 isolates reported in this study are all deposited and accessible at the EpiCoV database of GISAID (https://www.gisaid.org/). The corresponding GISAID accession codes are listed in Table 2.

## Funding Support

This study was partially funded by the Philippine Council for Health Research and Development, Department of Science and Technology Philippines and the University of the Philippines – Philippine General Hospital.

## Competing Interest

The SARS-CoV-2 isolates sequenced and reported in this study are part of the field validation done for the locally-developed GenAmplify nCoV rRT-PCR test kit.

## Data Availability

Genome sequences of the six Philippine SARS-CoV-2 isolates reported in this study are all deposited and accessible at the EpiCoV database of GISAID https://www.gisaid.org/). The corresponding GISAID accession codes are listed in **Table 2**.

## SUPPLEMENTARY MATERIALS

**Table S1.** List of variants observed across all six Philippine SARS-CoV-2 samples.

**Figure S1.** Overall sequence and structural similarity of the six Philippine SARS-CoV-2 scaffolds with respect to the NC_045512.2 reference sequence. The Mauve alignment shows a single sequence block (in red) for each of the scaffolds, suggesting highly identical genomic organizations. The sequences are also very similar at the nucleotide level, as shown by the red histogram inside the sequence blocks, except for a few breaks (depicted in white) that coincide with regions of ambiguous bases (N’s) in the scaffolds.

**Figure S2.** Partial amino acid sequence alignment of the ORF1ab gene RdRp region. The figure shows the alignment for residues 1-100, 401-600, and 801-900 of the RdRp region, containing all observed mismatches (total alignment length = 932 a.a.). The coverage (cov), percent identities (pid), and highlighted mismatches are relative to the NC_045512.2 reference sequence.

**Figure S3.** Partial amino acid sequence alignment of the spike glycoprotein (S) gene. The figure shows the alignment for residues 701-800 amino acids of the S gene product, containing the single observed mismatch (total alignment length = 1,274 a.a.). The coverage (cov), percent identities (pid), and highlighted mismatch are relative to the NC_045512.2 reference sequence.

**Figure S4.** Amino acid sequence alignment of the membrane glycoprotein (M) gene (total alignment length = 235 a.a.). The coverage (cov), percent identities (pid), and highlighted mismatches are relative to the NC_045512.2 reference sequence. The symbol X corresponds to ambiguous bases (N’s) or gaps in the underlying nucleotide sequence. The stretch of mismatched residues from positions 192-202 are the result of a 35 bp repeat in the nucleotide sequence assembly for sample PGC002.

**Figure S5.** Amino acid sequence alignment of the envelope protein (E) gene (total alignment length = 76 a.a.). The coverage (cov) and percent identities (pid) shown are relative to the NC_045512.2 reference sequence. No mismatches were observed.

**Figure S6.** Amino acid sequence alignment of the nucleocapsid phosphoprotein (N) gene (total alignment length = 420 a.a.). The coverage (cov), percent identities (pid), and highlighted mismatches are relative to the NC_045512.2 reference sequence.

## Notes

### Author Declarations

The study received ethics approval from the University of the Philippines Manila Research Ethics Board with approval number 2020-96 187-01.

